# Utility of thromboelastography to assess the effect of anticoagulation reversal in intracranial hemorrhage

**DOI:** 10.1101/2024.08.07.24311652

**Authors:** A Zepeski, BA Faine, M Ghannam, HM Olalde, L Wendt, A Naidech, NM Mohr, EC Leira

## Abstract

**Background:** Intracranial hemorrhage (ICH) is a serious complication associated with oral anticoagulant use and is associated with significant morbidity and mortality. Although anticoagulation reversal agents are utilized as standard of care, practitioners are limited in their ability to assess degree of anticoagulation reversal for direct oral anticoagulants (DOACs). There is a clinical need identify biomarkers to assess anticoagulation status in patients with DOAC-associated ICH to ensure hemostatic efficacy of anticoagulation reversal agents in the acute setting. The purpose of this study was to assess the utility of thromboelastography (TEG) to assess the impact of anticoagulation reversal in patients presenting to the emergency department (ED) with DOAC-associated ICH.

**Methods:** We conducted a prospective, observational cohort study in adult patients presenting to the ED with acute DOAC-associated ICH. Patients were excluded if last DOAC dose was >48 hours prior to hospital arrival, if they experienced polytrauma, were pregnant, incarcerated, had a history of hepatic failure or coagulopathy, or received anticoagulation reversal with products other than prothrombin complex concentrates (PCCs). We collected baseline TEG samples from participants prior to anticoagulation reversal, as well as 30-minutes, 12-hours, and 24-hours post-reversal. TEG samples were also collected from participants who transferred to our facility after reversal at ED presentation, as well as 12- and 24-hours post-reversal.

**Results:** Pre-reversal TEG was collected on 10 participants prior to DOAC reversal. A significant decrease in TEG R-time was observed at 30 minutes post-reversal. R-time increased at 12- and 24-hours to baseline levels. Significant changes were not observed in K-time, clot strength, maximum amplitude, or coagulation index.

**Conclusions:** TEG R-time may be able to detect a change in anticoagulation activity of DOACs in ICH after anticoagulation reversal. R-time decreases acutely after anticoagulation reversal and rebounds at 12- and 24-hours post-reversal.

## Introduction

Direct oral anticoagulants (DOACs), including factor Xa inhibitors have become the cornerstone of outpatient anticoagulation therapy for stroke prevention in atrial fibrillation and treatment of venous thromboembolism.^1,2^ Despite having a more favorable safety profile, factor Xa inhibitors still carry risk of bleeding complications, including intracranial hemorrhage (ICH). Use of oral anticoagulation has been shown to increase both short and long-term mortality in acute spontaneous and traumatic ICH.^3-5^ Acute spontaneous ICH is associated with 2 times greater mortality than ischemic stroke.^6^ Coagulation factor Xa [recombinant], inactivated-zhzo is recommended by guidelines to emergently reverse anticoagulation activity of apixaban or rivaroxaban, however prothrombin complex concentrates (PCCs) are frequently utilized with comparable efficacy and reduced thrombotic effects.^7-10^ Regardless of the reversal agent of Xa inhibitors that are used, clinicians are limited in current clinical practice by the lack of simple and reliable biomarkers to monitor anticoagulation activity in these patients.

Previous studies have outlined shortcomings of traditional coagulation assays to assess DOAC activity.^11^ Apixaban has no significant impact on activated partial thromboplastin time (aPTT), prothrombin time (PT), or international normalized ratio (INR). Rivaroxaban may increase PT/INR and aPTT, but only at high serum concentrations. The Annexa-4 Trial utilized anti-factor Xa assays as a surrogate to measure recombinant coagulation factor Xa activity.^12^ The use of Anti Xa levels remains limited in most emergent settings, although it has been correlated with hematoma expansion.^9,13^ Unfortunately, our ability to measure both the degree of anticoagulation activity and reversal in patients with acute DOAC-related bleed remains limited, which impacts optimal patient managment.^7,14-16^

Thromboelastography (TEG) is a coagulation study that has been utilized in trauma and surgical populations to provide a comprehensive, objective measurement of the combined impact of clotting factors, fibrin, fibrinogen, and platelet activity.^17^ Novel approaches of TEG application have investigated its utility in the emergency department (ED) to identify patients who have taken DOACs in the setting of bleeding or may need hemostatic treatment, particularly prior to neurosurgery.^18-20^ However, a significant evidence gap remains regarding the utility of TEG to measure the degree of anticoagulation reversal after pre-ICH anticoagulant reversal. The primary objective of our study is to determine the changes in TEG with anticoagulation reversal in patients with DOAC-associated ICH.

## Methods

This was a prospective, cohort study of patients presenting to a Level I Trauma Center/Comprehensive Stroke Center with acute ICH from November 2022 – August 2023. Patients were included in the study if they were taking rivaroxaban or apixaban, were 18 years of age or older, and had confirmed ICH on computed tomography (CT) imaging. Patients were excluded from enrollment if the last dose of DOAC was ≥ 48 hours prior to enrollment, if DOAC use could not be confirmed, if they were pregnant, incarcerated, presented with polytrauma (any identified injury other than ICH), had a history of hepatic failure or coagulopathy, or received coagulation factor Xa [recombinant], inactivated-zhzo for DOAC reversal. Patients with imminent death, receiving active cardiopulmonary resuscitation or transition to palliative care in the ED were also excluded from the study. Informed written consent was obtained by all participants or their legally authorized representative. The study was approved by the local Institutional Review Board. This study is reported in accordance with the STROBE publication criteria for observational studies.^21^

Participants were divided into 3 subgroups based on timing of PCC administration and TEG serum sampling. Administration of activated PCC was not an intervention of the study— data collection was completely observational, and tracked the biomarker response to anticoagulation reversal.

1. Group 1 included participants who received activated PCC for anticoagulation reversal at the enrolling institution with baseline TEG drawn prior to reversal as well at 0.5 hours, 12 hours, and 24 hours post-reversal.
2. Group 2 included participants who received anticoagulation reversal prior to TEG collection and had levels drawn after reversal (first available), 12 hours, and 24 hours post-reversal.
3. Group 3 included participants who were consented to the study but ultimately never received anticoagulation reversal. Group 3 participants had TEG samples drawn at time of consent (first available), 12 hours after ED presentation and 24 hours after ED presentation.

After patients consented to participate in the study, we collected whole blood samples in light blue sodium citrate tubes (a minimum volume of 2.7 mL of whole blood was required). Study staff delivered samples immediately to the core lab after collection.

Thromboelastography was performed on whole blood samples using TEG® 5000 hemostasis analyzer (Haemonetics®; Boston, MA) upon receipt in the laboratory.

Data collection included gender, age, admission creatinine clearance, admission Glasgow Coma Scale (GCS), admission National Institutes of Health (NIH) Stroke Scale, ICH location and volume, DOAC dose and indication, concomitant anticoagulants, medications used for anticoagulation reversal and dose, baseline anti-Xa assays, PTT, INR, and discharge disposition.

## Data Analysis

Repeated data (e.g., TEG values over time repeated for individual participants) were analyzed with linear mixed models using R, version 4.3.3 (R Project for Statistical Analysis, Vienna, Austria, 2004). For each outcome of interest, a univariate model was fit with timepoint as the predictor, analyzed categorically. The baseline timepoint was used as the reference group for comparisons and coefficients were generated for each subsequent timepoint along with 95% confidence intervals and p-values. Additionally, F-Tests from these models were used to assess the overall impact of Timepoint on the outcome of interest. Pearson’s correlation was also used to assess the relationship between baseline anti-Xa levels and R-time in the group 1 cohort, and the correlation coefficient and its corresponding p-value were reported. REDCap™ was used to securely store abstracted data (NIH and CTSA grant number: UM1TR004403).

## Results

A total of 117 patients were screened. Eighty were found to be non-eligible, and the reasons for exclusion are shown in Figure 1. Thirty-seven participants were enrolled in the study. Three of these patients were excluded due to protocol deviations, so 34 participants were included in the final analysis. Baseline characteristics of participants are included in Table 1. Ten participants (29%) were enrolled prior to administration of DOAC reversal (group 1), 21 participants (62%) received anticoagulation reversal prior to enrollment due to interfacility transfer (group 2). Three participants were enrolled in the study but did not receive DOAC reversal (group 3). The baseline characteristics of these patients is shown in table 1. Most participants were taking apixaban (n= 25, 74%) prior to arrival and the most common indication for DOAC use was stroke prevention in atrial fibrillation (n=27, 79%). Two-thirds of participants experienced ICH of traumatic etiology. Concomitant antiplatelet use was common with 38% of participants taking aspirin prior to arrival.

**Table 1.** Baseline Characteristics.

|  | Participants (n=34) |
| --- | --- |
| Age (years), median (IQR) | 81 (72-84) |
| Gender (male), (%) | 24 (70.6) |
| CrCl (mL/min), (%) |  |
| <30 | 4 (11.7) |
| 30-59 | 13 (38.2) |
| ≥60 | 17 (50.0) |
| Pre-admission DOAC (%) |  |
| Apixaban | 25 (73.5) |
| Rivaroxaban | 9 (26.5) |
| Time since last known well (hours), median (IQR)<sup>1</sup> |  |
| DOAC indication (%) |  |
| Atrial fibrillation | 27 (79.4) |
| Venous thromboembolism | 7 (20.6) |
| *Other | 3 (8.8) |
| Concomitant medications (%) |  |
| Aspirin | 13 (38.2) |
| Clopidogrel | 2 (5.9) |
| None | 20 (58.8) |
| ICH Type (%) |  |
| Traumatic | 23 (67.6) |
| Spontaneous | 10 (29.4) |
| Unknown | 1 (2.9) |
| TEG Sampling Group (%) |  |
| Group 1 | 10 (29.4) |
| Group 2 | 21 (61.7) |
| Group 3 | 3 (8.8) |
| Transfer Type (%) |  |
| Transfer from other facility | 26 (76.5) |
| Direct presentation | 8 (23.5) |
| Group 1 Characteristics |  |
| Time from last DOAC dose to study enrollment, median (IQR) | 11.1 (8.9-13.9) |
| Anti-Xa level (ng/mL), median (IQR) |  |
| Apixaban assay (n=8) | 158 (75-252) |
| Rivaroxaban assay (n=1) | 158 |
| activated PCC dose (units/kg), median (IQR) | 24.8 (20.5-29.7) |
| <sup>1</sup>Time from reported last known well to hospital presentation |  |
| <sup>2</sup>peripheral vascular disease (n=1), apical akinesis (n=1), transcatheter aortic valve replacement (n=1) |  |

**Figure 1.**
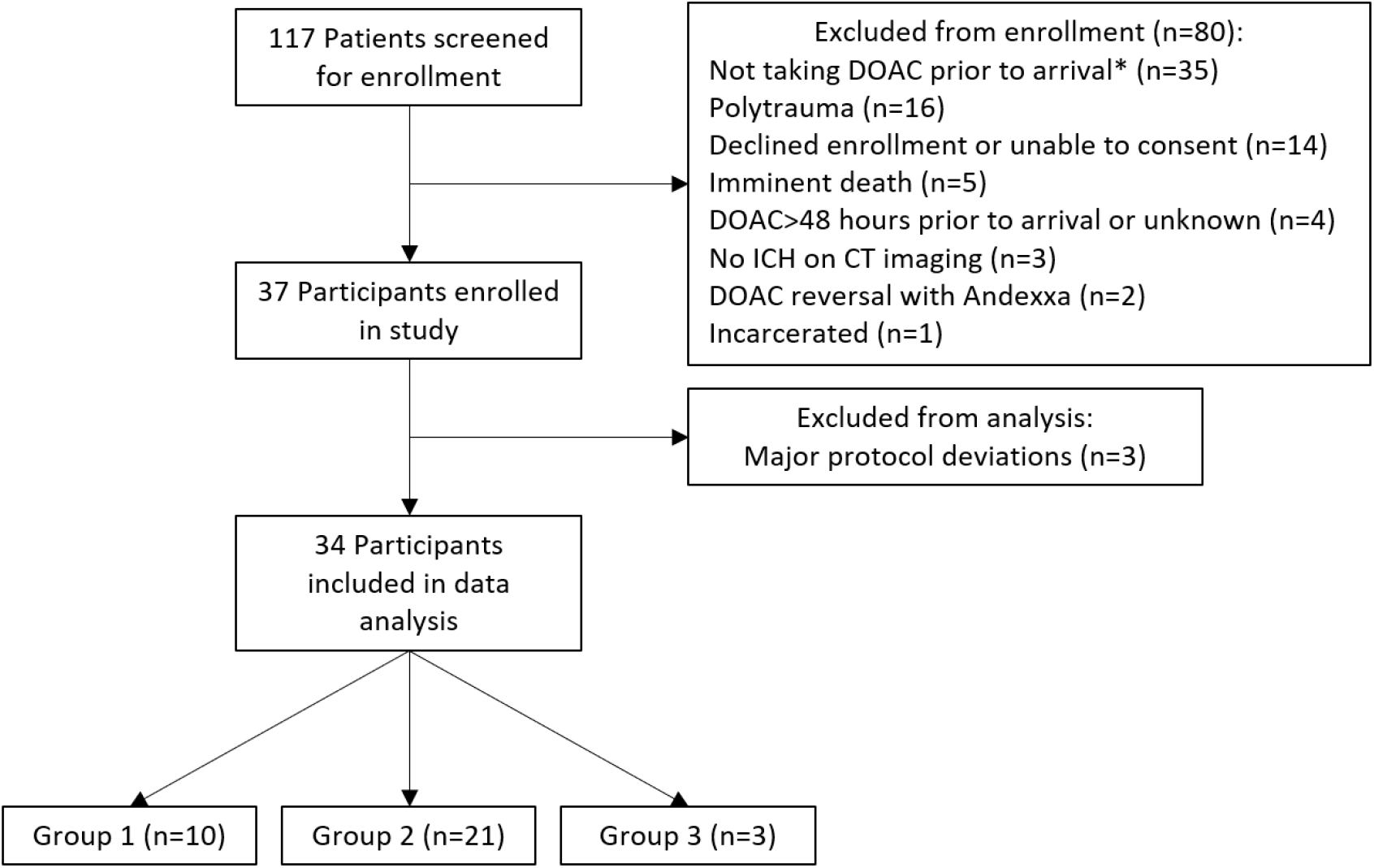
Participant screening and enrollment. Group 1: received activated PCC for anticoagulation reversal with baseline TEG drawn prior to reversal. Group 2: received anticoagulation reversal prior to TEG collection and had levels drawn after reversal. Group 3: consented to the study but never received anticoagulation reversal.

Ten participants were enrolled prior to anticoagulation reversal with PCC, all of whom received activated PCC based on institutional protocol. The median dose of activated PCC was 24.8 units/kg (IQR 20.5-29.7). Anti-Xa levels were drawn prior to anticoagulation reversal, which are reported in Table 1. Figure 2 illustrates TEG R-time values at baseline, 30-minutes post PCC reversal (group 1) and at 12 hours post-PCC reversal. Table 2 summarizes TEG parameters before and after anticoagulation reversal. There was not a statistically significant linear correlation between baseline anti-Xa levels and R-time in the group 1 cohort (R = -0.273, p = 0.417). The median R-time prior to anticoagulation reversal was 5.40 seconds (IQR 4.38-5.73) and 4.20 seconds (IQR 3.68-4.75) after reversal with activated PCCs. By 24 hours after reversal, R-time increased to 5.30 seconds (IQR 5.00-5.80). R-time values were shown to significantly decrease after DOAC reversal at 30 minutes (Beta = -0.91, p = 0.035), but were not significantly different at 12 and 24 hours compared to baseline values (Table 3). This effect was not observed when assessing clot Angle, or rate of clot formation.

**Table 2.** TEG Values Relative to Anticoagulation Reversal Administration (Group 1)

| **Characteristic Median (IQR)** | **Baseline, N = 10** | **30 Minutes post-PCC, N = 8<sup>2</sup>** | **12 Hours post-PCC, N = 10** | **24 Hours post-PCC, N = 9<sup>3</sup>** | **p-value<sup>2</sup>** |
| --- | --- | --- | --- | --- | --- |
| R-Time | 5.40<br>(4.38, 5.73) | 4.20<br>(3.68, 4.75) | 5.25<br>(4.43, 5.80) | 5.30<br>(5.00, 5.80) | **0.044** |
| K-Time | 1.25<br>(1.10, 1.30) | 1.05<br>(0.80, 1.23) | 1.20<br>(1.03, 1.38) | 1.20<br>(1.10, 1.30) | 0.999 |
| LY30 | 0.10<br>(0.03, 0.35) | 0.75<br>(0.00, 1.15) | 0.35<br>(0.05, 0.90) | 0.10<br>(0.00, 0.70) | **0.039** |
| Clot Strength | 13.70<br>(12.28, 14.23) | 10.70<br>(9.73, 12.45) | 13.75<br>(12.03, 14.73) | 12.70<br>(10.30, 15.30) | 0.309 |
| MA | 73.4<br>(71.0, 74.0) | 68.2<br>(66.0, 71.3) | 73.4<br>(70.7, 74.7) | 71.8<br>(67.3, 75.4) | 0.342 |
| CI | 3.10<br>(1.83, 3.58) | 3.35<br>(1.83, 3.83) | 2.30<br>(1.40, 3.35) | 2.60<br>(2.30, 3.40) | 0.736 |
| <sup>1</sup>F-Test from Linear Mixed Model |  |  |  |  |  |
| <sup>2</sup>Missing data are attributable to missed lab draw time or laboratory error (unable to run sample). |  |  |  |  |  |

**Table 3.**
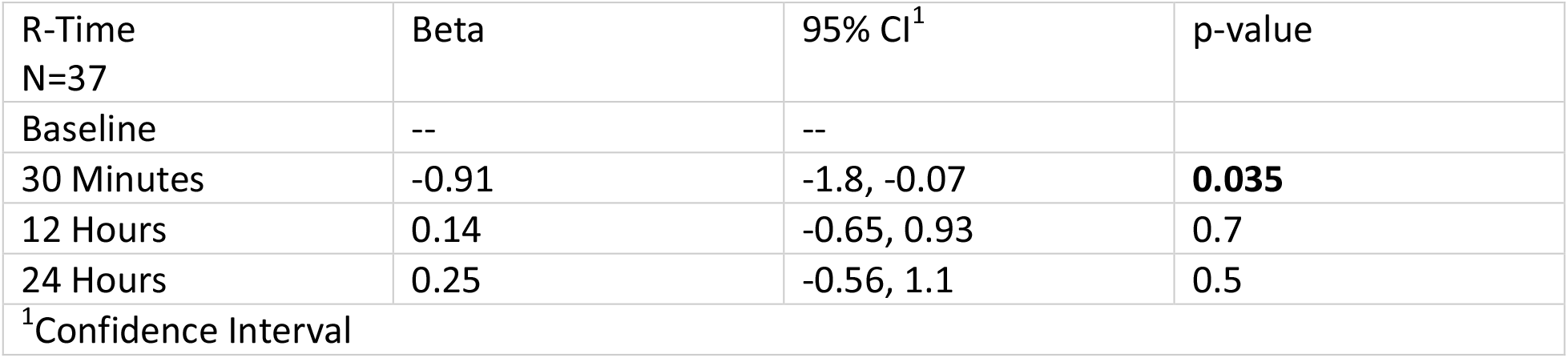
Pairwise Time Differences for TEG R-time (Group 1)

**Figure 2.**
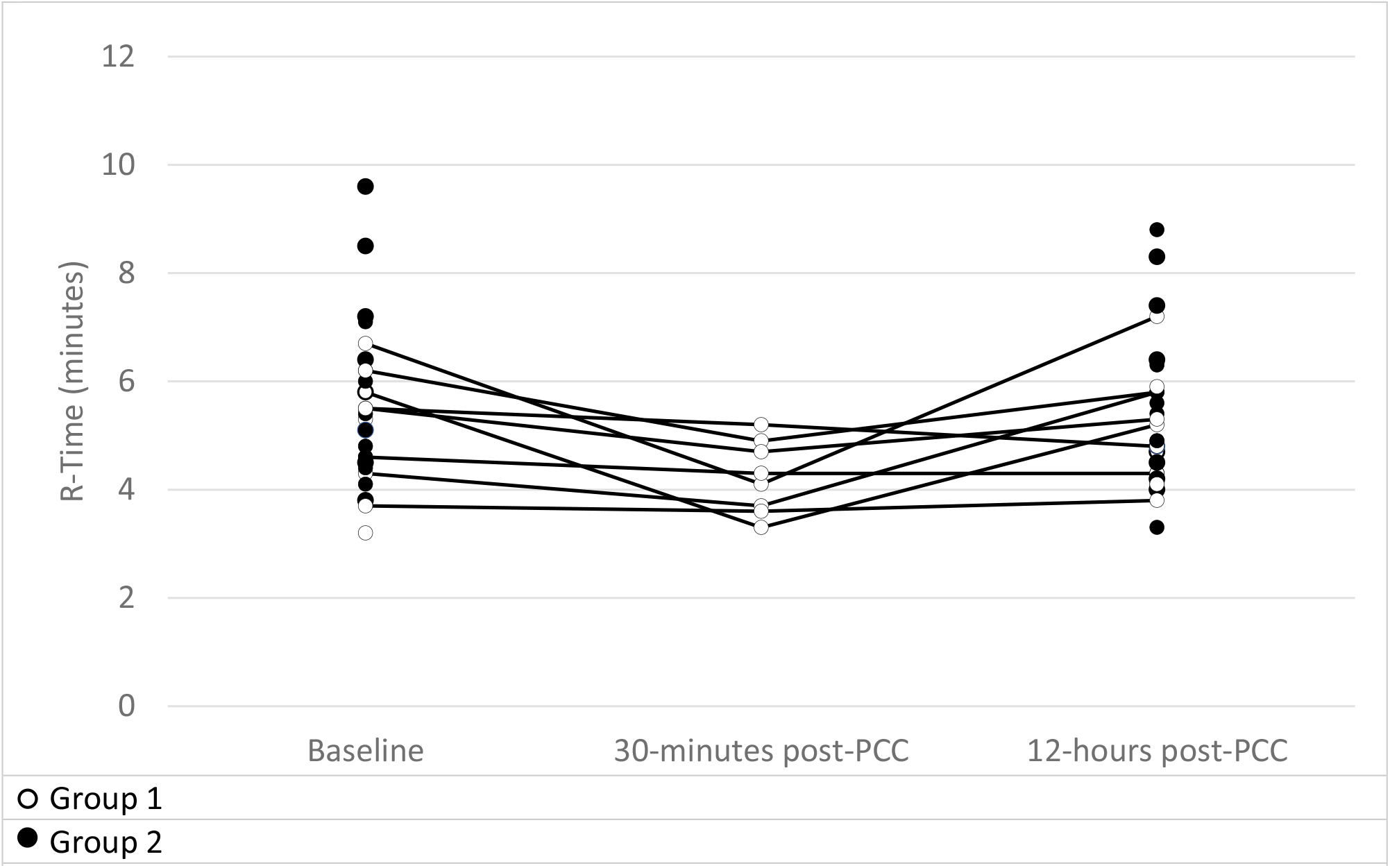
Change in TEG R-time over time (Groups 1 and 2) Baseline R-time was collected in Group 1 and 2 participants. Baseline was defined in group 2 participants as first available TEG upon arrival from outside hospital. 30-minute post-PCC TEG was only drawn in Group 1.

## Discussion

This was an exploratory study in real-world patients presenting to the ED with acute ICH. We prospectively measured TEG before and after the administration of activated PCC in patients with DOAC-associated ICH. We observed substantial changes in TEG parameters 30 minutes after treatment, with subsequent return to baseline measures. These results show that TEG is a potential biomarker to measure the effect of anticoagulant reversal in patients with DOAC-associated ICH. These results suggest TEG may be beneficial in addition to measures of anti-Xa activity in patients with DOAC-associated ICH. Anti-factor Xa assays have been utilized in the ED setting to assess presence of DOAC but the defined utility in determining degree of anticoagulation remains limited. Our findings suggest that rivaroxaban and apixaban assays were not associated with baseline R-time prior to reversal. This may further support the limitations of anti-Xa assays in determining degree of anticoagulation in patients requiring emergent DOAC reversal, despite their specificity for the presence of factor Xa inhibitors.

Although all participants were treated with DOACs prior to ICH, variability in baseline R-time values DOAC-associated ICH was observed. This is not an unexpected result as apixaban and rivaroxaban do not rely on standardized measured levels of anticoagulation for observed efficacy.^22,23^ Other studies have utilized DOAC-calibrated TEG 6s to identify presence of DOAC, utilizing specific R-time thresholds, but have not assessed changes to R-time after administration of anticoagulation reversal.^20^ Despite the observed variability, R-time significantly decreased after activated PCC administration, underscoring improvement in coagulation with activated PCC administration. A change in R-time may be a more valuable marker of anticoagulation reversal and hemostasis rather than achievement of an absolute R-time value.

R-time increased as measured at 12- and 24-hour intervals after the administration of PCCs. This rebound in R-time is not unexpected and is consistent with the estimated 8-12 hour duration of action of PCCs.^24^ It is for this same rationale, that guidelines recommend concomitant administration of vitamin K with PCCs to ensure sustained reversal of warfarin anticoagulant activity.^10^ Overall, these trends in R-time relative to anticoagulation reversal with PCCs are expected as R-time reflects time to beginning of clot formation, which is driven by coagulation factors. Artang et al. reported TEG R-time was significantly and positively correlated with measured DOAC concentrations in healthy volunteers. The same association was not found in alpha-angle (surrogate for fibrinogen activity), and maximum amplitude (surrogate for platelet activity).^18^ The observed changes in TEG biomarkers after reversal suggest that this technique may be useful to guide this treatment. Future analyses might test the hypothesis that changes in TEG biomarkers are predictive of hematoma expansion as measured with semi-automated voxel-based techniques, a modifiable predictor of worse outcome in patients with acute ICH.

We recognize the limitations of our study. First, there was a high level of participant heterogeneity in the sample and notably a mix of traumatic and spontaneous ICH patients. Trauma-induced coagulopathy can lead to changes in TEG values and may confound interpretation of TEG in patients on anticoagulation, especially measurements of LY30.^25^ It can be difficult to determine etiology of ICH acutely in the absence of known severe trauma, therefore all ICH was intentionally included in the sample and etiology was determined based on chart review after hospital discharge. Additionally, the investigators attempted to address this the impact of trauma-induced coagulopathy by excluding patients with polytrauma.

Second, several of the participants received anticoagulation reversal prior to enrollment, limiting the proportion of the sample with baseline TEG values prior to reversal. Because of the nature of our referral center, a sizeable number of patients are transferred from outside hospitals which limits our ability to obtain pre-treatment baseline levels.

## Conclusion

In patients with DOAC-associated ICH, the administration of activated PCC was associated changes in TEG biomarkers of hemostasis at 30 minutes that returned to baseline at 12 hours. Future research might determine if these changes predict hematoma expansion and determine biomarkers of efficacy for anticoagulant reversal.

## Data Availability

All data referenced in this manuscript are available and stored securely in the University of Iowa REDCap database.

## Acknowledgments

None

